# Polygenic effects on the risk of Alzheimer’s disease in the Japanese population

**DOI:** 10.1101/2023.10.06.23296656

**Authors:** Masataka Kikuchi, Akinori Miyashita, Norikazu Hara, Kensaku Kasuga, Yuko Saito, Shigeo Murayama, Akiyoshi Kakita, Hiroyasu Akatsu, Kouichi Ozaki, Shumpei Niida, Ryozo Kuwano, Takeshi Iwatsubo, Akihiro Nakaya, Takeshi Ikeuchi, Alzheimer’s Disease Neuroimaging Initiative, the Japanese Alzheimer’s Disease Neuroimaging Initiative

**Affiliations:** Department of Computational Biology and Medical Sciences, Graduate School of Frontier Science, The University of Tokyo, Chiba, Japan; Department of Medical Informatics, Graduate School of Medicine, Osaka University, Osaka, Japan; Department of Molecular Genetics, Brain Research Institute, Niigata University, Niigata, Japan; Brain Bank for Aging Research (Department of Neuropathology), Tokyo Metropolitan Institute of Geriatrics and Gerontology, Tokyo, Japan; Brain Bank for Neurodevelopmental, Neurological and Psychiatric Disorders, United Graduate School of Child Development, Osaka University, Osaka, Japan; Department of Pathology, Brain Research Institute, Niigata University, Niigata, Japan; Department of General Medicine & General Internal Medicine, Nagoya City University Graduate School of Medicine, Nagoya, Japan; Medical Genome Center, Research Institute, National Center for Geriatrics and Gerontology, Aichi, Japan; RIKEN Center for Integrative Medical Sciences, Kanagawa, Japan; Core Facility Administration, Research Institute, National Center for Geriatrics and Gerontology, Aichi, Japan; Asahigawaso Research Institute, Social Welfare Corporation Asahigawaso, Okayama, Japan; Department of Neuropathology, Graduate School of Medicine, The University of Tokyo, Tokyo, Japan

**Keywords:** Polygenic Risk Score, Alzheimer’s Disease, Mild Cognitive Impairment

## Abstract

**Background:** Polygenic effects have been proposed to account for some disease phenotypes; these effects are calculated as a polygenic risk score (PRS). This score is correlated with Alzheimer’s disease (AD)-related phenotypes, such as biomarker abnormalities and brain atrophy, and is associated with conversion from mild cognitive impairment (MCI) to AD. However, the AD PRS has been examined mainly in Europeans, and owing to differences in genetic structure and lifestyle, it is unclear whether the same relationships between the PRS and AD-related phenotypes exist in non-European populations. In this study, we calculated and evaluated the AD PRS in Japanese individuals using GWAS statistics from Europeans.

**Methods:** In this study, we calculated the AD PRS in 504 Japanese participants (145 cognitively unimpaired (CU) participants, 220 participants with late mild cognitive impairment (MCI), and 139 patients with mild AD dementia) enrolled in the Japanese Alzheimer’s Disease Neuroimaging Initiative (J-ADNI) project. In order to evaluate the clinical value of this score, we (1) determined the polygenic effects on AD in the J-ADNI and validated it using two independent cohorts (a Japanese neuropathology (NP) cohort (n=565) and the North American ADNI (NA-ADNI) cohort (n=617)), (2) examined the AD-related phenotypes associated with the PRS, and (3) tested whether the PRS helps predict the conversion of MCI to AD.

**Results:** The PRS using 131 SNPs had an effect independent of *APOE*. The PRS differentiated between CU participants and AD patients with an area under the curve (AUC) of 0.755 when combined with the *APOE* variants. Similar AUC was obtained when PRS calculated by the NP and NA-ADNI cohorts was applied. In MCI patients, the PRS was associated with cerebrospinal fluid phosphorylated-tau levels (β estimate = 0.235, p value = 0.026). MCI with a high PRS showed a significantly increased conversion to AD in *APOE* ε4 noncarriers with a hazard rate of 2.22.

**Conclusions:** We showed that the AD PRS is useful in the Japanese population, whose genetic structure is different from that of the European population. These findings suggest that the polygenicity of AD is partially common across ethnic differences.

## 1. Background

Alzheimer’s disease (AD) is a neurodegenerative disease caused by environmental and genetic factors [1, 2]. Environmental factors, which are acquired and modifiable, associated with AD include smoking status, alcohol consumption, diet, and physical activity [3]. On the other hand, the heritability of AD is approximately 70%, and genetic factors are inborn and nonmodifiable [4, 5]. However, knowing one’s genetic risk early in life can motivate one to improve modifiable factors. Indeed, sharing genetic test results with carriers of genetic risk for disease may promote behavioural changes rather than increase psychological distress [6, 7]. Thus, knowledge of the individual genetic risk of AD is expected to contribute to delaying the onset of AD and early therapeutic intervention.

The largest genetic risk factor for AD is the ε4 allele of the apolipoprotein E (*APOE*) gene, but *APOE* ε4 explains only approximately 10% of AD cases based on heritability [4, 5]. In addition, even when other AD-associated genetic variants found in previous genome-wide association studies (GWAS) are also considered, they do not explain all the genetic variance in AD patients [8], suggesting the existence of additional unknown AD-related genetic variants. To clarify this “missing heritability”, polygenic effects that aggregate the small effects of many alleles have been proposed to underlie AD.

Polygenic risk score (PRS) is a measure to quantify the combined effect of genetic variants on an individual’s risk for disease. The combination of the *APOE* ε4 allele dose and PRS has been shown to improve disease prediction accuracy in the European population [9]. Moreover, the PRS is associated with AD-related phenotypes, such as brain volumes [10–12], brain amyloid-beta (Aβ) burden [11, 12], and plasma phosphorylated tau [13], and has been reported to be useful in predicting conversion from mild cognitive impairment (MCI) to AD [14, 15].

However, the clinical application of the PRS must be approached with caution. One of several concerns is that the effects of the PRS are not consistent across races [16, 17]. This is because genetic structures, such as linkage disequilibrium (LD) blocks, are different across populations and because the GWAS summary statistics used as a weight for each single-nucleotide polymorphism (SNP) to calculate the PRS are based primarily on people of European ancestry. Taking a PRS calculation method based on GWAS summary statistics from European individuals and applying it to non-European individuals compromises prediction accuracy since the genetic risk of that population may not be reflected properly [18]. Therefore, for future clinical application of the AD PRS, it is necessary to evaluate the utility of this score in populations of different ancestry. In addition, harmonization of protocols such as inclusion and exclusion criteria is critical for rigorous comparisons between different cohorts.

Therefore, in this study, we calculated the AD PRS in 504 Japanese participants (145 cognitively unimpaired participants, 220 participants with late MCI, and 139 patients with mild AD dementia) enrolled in the Japanese Alzheimer’s Disease Neuroimaging Initiative (J-ADNI) project and evaluated its effectiveness in the North American ADNI (NA-ADNI) cohort including North American 1,070 participants. The J-ADNI study used a harmonized protocol to the NA-ADNI study. The previous comparative study of AD dementia between the US and Japan in the ADNI projects reported that MCI in the Japanese population shows similar progression profile as MCI in North America in terms of cognitive function [19]. We moreover validated the AD PRS using independent genomic data from 565 Japanese individuals with a neuropathological diagnosis by autopsy. Furthermore, we also examined the AD endophenotypes in association with PRS, and tested whether the PRS is useful for predicting conversion from MCI to AD.

## 2. Materials and methods

### 2.1 Japanese participants from the J-ADNI cohort

Data used in the preparation of this article were obtained from the J-ADNI database deposited in the National Bioscience Database Center Human Database, Japan (Research ID: hum0043.v1, 2016) [19]. This database enrolled cognitively unimpaired (CU) participants, participants with late MCI, and patients with mild AD dementia (ADD) using criteria consistent with those of the North American ADNI (NA-ADNI) [20]. The J-ADNI was launched in 2007 as a public–private partnership led by Principal Investigator Takeshi Iwatsubo, MD. The J-ADNI was aimed to test whether serial magnetic resonance imaging (MRI), positron emission tomography (PET), other biological markers, and clinical and neuropsychological assessment can be combined to measure the progression of late MCI and mild ADD in the Japanese population. The J-ADNI did not recruit participants with early MCI. The ethics committees of the University of Tokyo, Osaka University and Niigata University approved the study.

A total of 715 volunteer participants between the ages of 60 and 84 years were diagnosed with late MCI or mild ADD or were CU and considered for inclusion in the J-ADNI. Of the 715 participants assessed for study eligibility, 537 met the criteria and were enrolled. Of these 537 participants, 508 (CU, 147; MCI, 221; ADD, 140) underwent genotyping analysis. Participants were evaluated every 6 or 12 months over a period of 36 months for CU and MCI participants and over a period of 24 months for participants with ADD, as in the NA-ADNI. As detailed below, the J-ADNI collected various imaging, clinical and neuropsychological data from these participants in addition to the genomic data. These data were obtained from the database described above.

### 2.2 Japanese neuropathological cohort

An independent neuropathological (NP) cohort composed of 577 brain donors was used for PRS validation [21]. Of these donors, 365 control donors had little pathological findings associated with AD and 212 case donors had those consistent with AD. All ADD patients were neuropathologically diagnosed by senile plaque and neurofibrillary tangle. No neuropathological features of other neurodegenerative disorders such as dementia with Lewy body disease, frontotemporal lobal degeneration, and Parkinson’s disease, were observed. Control individuals did not show the typical neuropathological hallmarks of AD. As no clinical diagnosis is provided in this cohort, the term case or control is used in this study. As shown below, 565 brain donors (358 controls and 207 cases) passed QC. The demographic data of all the participants from the NP cohort are shown in **Table S1**.

### 2.3 Genotyping, quality control, and imputation

Whole blood samples from 508 participants in the J-ADNI cohort and post-mortem frontal cortices from 577 donors in the NP cohort were genotyped using the Infinium Asian Screening Array (Illumina), containing 657,490 SNPs. *APOE* genotypes in each participant were determined by haplotypes derived from rs7412 and rs429358, which were genotyped using TaqMan Assays (Applied Biosystems). We excluded SNPs that (i) had duplicated genomic positions, (ii) had low call rates (<5%), (iii) deviated from Hardy-Weinberg equilibrium compared to controls (*p* < 1×10^-5^), or (iv) had low minor allele frequency (<0.01). For QC purposes, we excluded participants who (i) had sex inconsistencies, (ii) had autosomal heterozygosity deviation (|*F_het_*| ≥ 0.2), (iii) had <99% of their genotypes called, or (iv) were in the same family according to pi-hat (>0.2). Furthermore, we used principal component analysis to remove outliers based on the 1000 Genomes Project samples [22]. Finally, 451,713 autosomal SNPs and the samples, including 504 participants from the J-ADNI cohort and 565 brain donors from the NP cohort passed the QC procedures.

Next, we performed phasing with Eagle v2.4.1 [23] and imputation with Minimac4 [24] using the whole-genome sequencing data of 3,541 participants obtained from the BioBank Japan Project [25] and the 1000 Genomes Project [22] as reference genome data. After repeating the above QC procedure for the imputed SNP markers, we excluded SNPs with poor imputation quality (*r^2^* ≤ 0.3). Finally, we obtained 7,633,670 SNPs and the samples, including the 504 participants from the J-ADNI (CU, 145; MCI, 220; and ADD, 139) and 565 brain donors from the NP cohort (control, 358; case, 207).

### 2.4 The NA-ADNI genetic data

The independent cohort data used in this study were obtained from the NA-ADNI [26]. The NA-ADNI was launched in 2003 as a public–private partnership led by Principal Investigator Michael W. Weiner, MD. The NA-ADNI was aimed to test whether serial MRI and PET data and the analysis of other biological markers and clinical and neuropsychological assessments can be combined to characterize the progression of MCI and early ADD.

SNP data from the NA-ADNI project were available for 1,674 participants across ADNI 1 and ADNI GO/2. Genotyping was conducted using three different platforms: Human610-Quad, HumanOmniExpress and Omni 2.5 M (Illumina) [27]. The SNP data were imputed using the TOPMeD imputation server after identical marker QC and sample QC as was used for the J-ADNI was performed. The SNP data analysed on each of the three platforms were imputed separately. After repeating the QC for the imputed SNP markers, we excluded SNPs with poor imputation quality (*r^2^* ≤ 0.3). If a participant was genotyped on more than one genotyping array, the dataset with the fewest missing values was selected.

According to the following procedures, we selected participants with predicted central European ancestry and self-reported white non-Hispanic ethnicity. For predicted ancestry, we used SNPweights software to infer genetic ancestry from genotyped SNPs [28]. The reference panel comprised European, West African, East Asian and Native American ancestral populations. Participants with predicted central European ancestry of 80% or more were retained. We obtained self-reported ethnicity information from the NA-ADNI database. The clinical diagnosis at the final visit was used to categorize the data. Furthermore, four participants who had significant memory concerns but no cognitive impairment were excluded. Finally, 1,482 participants (CU, 377; MCI, 481; and ADD, 624) remained.

Of the 1,482 participants, 412 participants were participants in the Alzheimer’s Disease Genetics Consortium (ADGC) and were included in the meta-analysis of AD GWAS used as SNP weights in the PRS calculation described below. We analysed a set of 1,070 participants (CU, 257; MCI, 453; and ADD, 360), excluding the 412 participants to avoid overfitting. The demographic data of all the participants from the NA-ADNI cohort are shown in **Table S2**.

### 2.5 Calculation of the PRS and prediction accuracy

The PRS was calculated for each individual and is expressed as the following weighted sum:

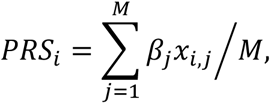

where *PRS_i_* is the PRS for individual *i*; *M* is the total number of SNPs used in the calculation; *β_j_* is the weight of *SNP_j_*, defined according to the effect size calculated by an independent GWAS; and *x_i,j_* is the number of minor alleles of *SNP_j_* that individual *i* has, thus has a value of 0, 1, or 2. In other words, the more minor alleles that are strongly associated with the disease, the higher the PRS.

SNPs included in the PRS were determined by the clumping and thresholding (C+T) method, the most common and supported method in AD studies [29, 30]. We used PRSice software implementing the C+T method to calculate the PRS [31]. The clumping method preferentially retains markers most strongly associated with disease from correlated markers in the same LD block. The thresholding method removes variants with GWAS p values greater than the selected p value threshold (*p_T_*) (*p* > *p_T_*). To determine the optimal *p_T_*, we tested *p_T_* values of 1×10^-6^, 1×10^-5^, 1×10^-4^, 1×10^-3^, 1×10^-2^, 0.05, 0.5, and 1.0. SNPs were weighted by their effect sizes (beta coefficient) from the AD GWAS in the European population [32].

The ability of the PRS to accurately classify CU participants and ADD patients was estimated in terms of (1) Nagelkerke’s *R^2^*, the proportion of the variance explained by the regression model, and (2) the area under the receiver operator characteristic curve (AUC). To calculate Nagelkerke’s *R^2^*, we constructed a logistic regression model, including the PRS and the first two components from the multidimensional scaling (MDS) analysis (full model), and compared it to a model with only the first two MDS components (null model). We assessed the difference in Nagelkerke’s *R^2^* between the full and null models (*R^2^* = *R^2^_Full_– R^2^_Null_*) and used the *p*_T_ corresponding to the highest value of Nagelkerke’s *R^2^*. The Nagelkerke’s *R^2^* was calculated by PRSice software [31]. The AUC was calculated based on the prediction results of the logistic regression model using the J-ADNI cohort as a test cohort. We also performed 5-fold cross validation (CV) to evaluate a predictive performance in a test cohort. We estimated the 95% credible intervals by using the ci.auc function from the R package “pROC”. DeLong’s test was conducted to assess potential significant differences between curves using the roc.test function from the R package “pROC”.

### 2.6 CSF biomarkers

In the J-ADNI cohort, cerebrospinal fluid (CSF) samples were assayed for Aβ(1–42), total tau (tTau), and phosphorylated tau (pTau) by using a multiplex xMAP Luminex platform (Luminex Corp, Austin, TX) with an Innogenetics (INNO-BIA AlzBio3; Ghent, Belgium) immunoassay kit-based reagent [33]. Of the 504 participants who underwent genotyping, 192 participants (CU, 52; MCI, 85; ADD, 55) also underwent CSF biomarker measurements at baseline.

### 2.7 Structural MRI and PET imaging

All participants in the J-ADNI cohort underwent a structural MRI scan at a signal strength of 1.5 tesla using a three-dimensional magnetization-prepared rapid-acquisition gradient-echo sequence according to a standardized protocol [34]. Cross-sectional and longitudinal processing streams in FreeSurfer, version 5.3, were used to estimate the atrophic changes in specific regions; we also evaluated the cortical thickness extracted in the longitudinal analysis. Of the 504 participants who underwent genotyping, the entorhinal cortex and hippocampus of 443 participants (CU, 133; MCI, 196; ADD, 114) was assessed by the FreeSurfer longitudinal stream. Each cortical thickness value was adjusted by the total intracranial volume.

Of the 504 participants, 315 and 162 individuals underwent a positron emission tomography (PET) scan using ^18^F-2-fluoro-2-deoxy-D-glucose (FDG) and ^11^C-Pittsburgh compound B (PiB), respectively. The PET scanning protocol was standardized to minimize the inter-site and inter-scanner variability [35]. All PET images went through the J-ADNI PET QC process [35]. The FDG PET images were classified into seven categories based on the criteria of Silverman *et al.* [36]. We analysed only PET images of 110 participants classified as having a normal pattern (N1 pattern) and 161 participants classified as having an AD pattern (P1 pattern). For PiB PET, the visual interpretation of four cortical areas on each side (frontal lobe, lateral temporal lobe, lateral parietal lobe, and precuneus/posterior cingulate gyrus) was evaluated by classifying PiB uptake in each cortical region as positive, equivocal, or negative. Cases with one or more positive cortical areas were defined as amyloid scan positive, and those with negative results in all four cortical regions were defined as amyloid scan negative. Other cases were considered equivocal. We analysed 65 negative and 87 positive amyloid scans, excluding 10 participants who were judged to be equivocal.

### 2.8 Neuropsychological tests

All participants in the J-ADNI cohort underwent the following neuropsychological tests: Mini–Mental State Examination (MMSE), Functional Assessment Questionnaire (FAQ), Clinical Dementia Rating Scale Sum of Boxes (CDR-SB), and AD Assessment Scale–Cognitive Subscale (ADAS-Cog).

### 2.9 Statistical analyses

Gene functional enrichment analysis of the closest genes around SNPs included in the PRS was performed using the Metascape database (http://metascape.org/) [37].

For the association analyses between the PRS and endophenotypes, we compared slopes with zero by linear regression model analyses. The covariates included age at baseline examination, sex, years of education, the first two principal components (PCs), and doses of *APOE* ε4 and ε2 alleles. P values were adjusted by false discovery rate (FDR) to avoid type I error.

Cox proportional hazards models using months of follow-up as a time scale were used to analyse the effects of PRSs on incident AD, presented as hazard ratios (HRs) and 95% confidence intervals (CIs) derived from a model with the following covariates: age at baseline examination, sex, years of education, the first two PCs, and dose of *APOE* ε4 and ε2 alleles. We analysed 208 MCI participants over a follow-up period of ≥ 12 months. Nonconverters were censored at the end of follow-up. Log-rank test was performed to examine the difference in conversion to AD between two PRS groups. This test was performed using only the PRS without covariates because the covariates other than PRS could affect the differences between the groups. Cox proportional hazard model analyses and log-rank tests were performed using the coxph and survdiff functions from the R package “survival”, respectively.

## 3. Results

### 3.1 The PRS successfully distinguish ADD patients and CU individuals in the J-ADNI cohort

After quality control of the genotyping data, the J-ADNI cohort included the 504 participants. The group with ADD had a higher mean age (p value <.001), a lower mean length of education (p value <.001), and a higher frequency of *APOE* ɛ4 carriers (p value <.001) than the CU group, whereas no differences were found in sex (p value = 0.429) or the frequency of *APOE* ɛ2 carriers (p value = 0.292) (**Table 1**).

We investigated whether the PRSs that were calculated using the statistics from the AD GWAS in the European population [32] are useful for discriminating between patients with ADD and CU individuals in the Japanese population. We calculated PRSs for 145 CU participants and 139 patients with ADD from the J-ADNI cohort. Our model using 173 SNPs showed the highest predictive power at *p_T_* < 1×10^-5^ and had a Nagelkerke’s *R^2^* of 0.167 (**left side of Table 2**), indicating that it explained more than 15% of the variance between the CU and ADD groups.

Given the known predictive power of SNPs in the *APOE* region for AD, we next removed this region from our PRS calculation to evaluate the predictive power of other loci. To exclude the effect of *APOE*, we excluded ±500 kb around *APOE* (**Figure S1**). This PRS, referred to as the PRS.noAPOE, was used in subsequent analyses. The predictive power of the PRS.noAPOE was the highest for *p_T_* < 1×10^-5^, with a Nagelkerke’s *R^2^* of 0.085 (**right side of Table 2**). The normalized values of the PRS.noAPOE of the ADD patients were significantly higher than those of the CU and MCI participants (p value < 0.05, Tukey’s honestly significant difference (HSD) test; **Figure 1**), while there was no significant difference between the CU and MCI participants (p value = 0.180, Tukey’s HSD test; **Figure 1**). These results suggest that the PRS contribute to distinguish between ADD patients and CU individuals in J-ADNI cohort even when the *APOE* region is excluded.

**Figure 1.**
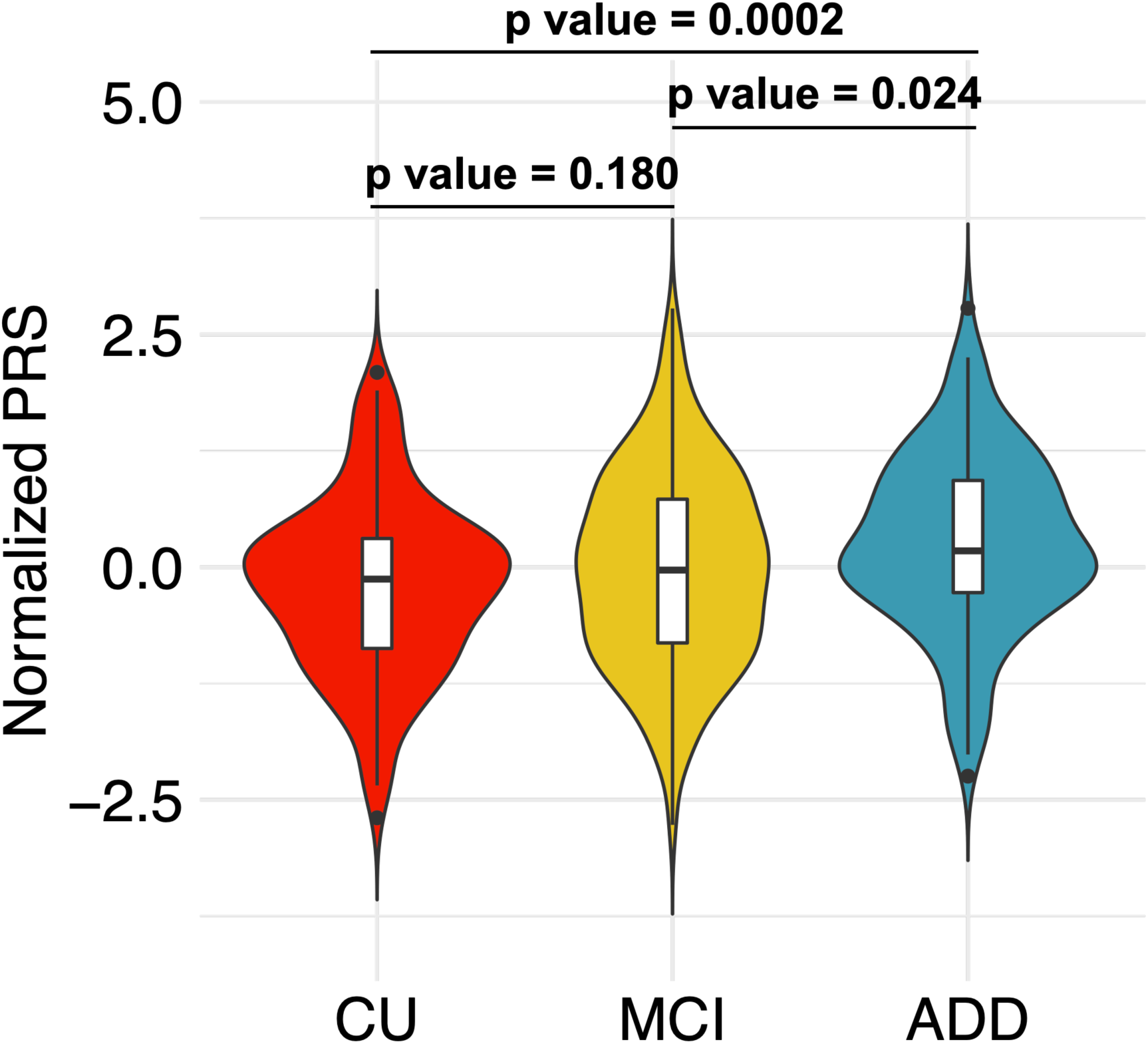
The PRS.noAPOE in the ADD group was significantly higher than those in the CU and MCI groups. The PRS.noAPOEs in each group were represented by violin plots (CU, n=145; MCI, n=220; ADD, n=139). Each violin plot includes the kernel probability density of the data at different values and the box plots with the median value and the interquartile range. Tukey’s HSD test was used to perform multiple comparisons of PRSs among each group. We normalized the PRS distribution to have a mean of 0 and an SD of 1. CN = cognitively normal; MCI = mild cognitive impairment; ADD = Alzheimer’s disease dementia.

### 3.2 The PRS in combination with the *APOE* alleles improves predictive power

Next, we examined whether the PRS.noAPOE and the characteristics of the participants independently influence the predictive power in J-ADNI cohort. The PRS.noAPOE was not correlated with sex, years of education, age at baseline examination, or the dose of the *APOE* ε4 or ε2 allele, even when participants were stratified into CU, MCI, and ADD groups (p value > 0.05; **Figure S2**). These results suggest that these factors contribute independently to the discrimination of AD and that combinations of these factors improve discrimination accuracy. We constructed a model including only the PRS.noAPOE and doses of *APOE* ε4 and ε2 alleles. This model showed predictive performance of AUC = 0.755 (95% CI = 0.695-0.807) (**Table 3**). The predictive performance of a monogenic model of only *APOE* alleles without the PRS.noAPOE was AUC = 0.696 (95% CI = 0.640-0.751) (**Table 3**). The addition of polygenic effects significantly improved the predictive accuracy of the monogenic model using only *APOE* (p value = 9.36×10^-4^, DeLong test). Additionally, the PRS model incorporating *APOE* alleles independently (PRS.noAPOE + *APOE* doses) has higher accuracy than the PRS model that includes SNPs in the *APOE* region (PRS.incAPOE) (AUC = 0.706; 95% CI = 0.643-0.764; p value = 0.049, DeLong test). Therefore, we constructed a predictive model including the PRS.noAPOE, sex, years of education, age at baseline examination, and doses of *APOE* ε4 and ε2 alleles. This model showed discriminative performance of AUC = 0.855 in distinguishing between the ADD patients and CU individuals in the J-ADNI cohort (95% CI = 0.808-0.898) (**Table 3**). These predictive performances showed the similar tendencies when evaluated by 5-fold CV (**Table S3**). Taken together, these results showed that the PRS based on European GWAS statistics was useful in discriminating between patients with ADD and CU participants in the Japanese population. Furthermore, the PRS had an effect independent of *APOE* alleles, and their combination improved predictive accuracy.

### 3.3 The effect of our PRS model is replicated in the independent cohorts

To examine the predictive accuracy of PRS.noAPOE in independent cohorts, we calculated the PRS values for 565 brain donors in the NP cohort (control, 358; case, 207) and 617 participants (CU, 257; ADD, 360) in the NA-ADNI using our PRS.noAPOE model. We note that the samples from the NP cohort received a definitive diagnosis based on the typical neuropathological hallmarks of AD using autopsy brains. The logistic regression model constructed in the J-ADNI cohort was applied to each cohort to assess discrimination accuracy. The predictive performance of PRS.noAPOE for the NP cohort was lower than that for the J-ADNI cohort (AUC = 0.550 (95% CI = 0.500-0.599)), but when *APOE* alleles were added, the predictive performance was replicated (AUC = 0.731 (95% CI = 0.686-0.773)) (**Table 3**).

In the NA-ADNI cohort, the imputed genotyping data included 130 of the 131 SNPs used in the PRS.noAPOE. A similar analysis in the NA-ADNI cohort also showed that the predictive performance of PRS.noAPOE in combination with *APOE* alleles (AUC = 0.730 (95% CI = 0.692-0.767)) was similar to that of the NP cohort. These analyses showed the reproducibility of our PRS model in independent cohorts.

### 3.4 ADD in the J-ADNI shows the polygenicity related to immune pathway

In order to examine the polygenicity of our PRS, we compared a model including only the PRS.noAPOE with a single-variable model for each of the 131 SNPs comprising the PRS.noAPOE. The single models with individual SNPs showed AUCs of 0.499 to 0.605 (median AUC = 0.515), while the model including only the PRS.noAPOE showed an AUC of 0.640 (95% CI = 0.576-0.704) (**Tables 3 and S4**), suggesting that the PRS.noAPOE reflects a polygenic effect. Here, SNPs with AUCs of less than 0.5 indicate protection rather than risk in our data.

We examined the genes closest to 131 SNPs included in the PRS.noAPOE. We found the 97 closest genes located within ±100 kb around the SNPs. These genes were associated with leukocyte-mediated immunity (FDR = 3.78×10^-5^), haematopoietic cell lineage (FDR = 4.45×10^-5^), the amyloid precursor protein (APP) catabolic process (FDR = 5.16×10^-5^), regulation of transferase activity (FDR = 3.57×10^-4^), and glial cell proliferation (FDR = 5.60×10^-3^) (**Table S5**). Overall, we found that the integrated scores of multiple SNPs around genes mainly associated with immune pathways may explain the Japanese AD traits.

### 3.5 The PRS associates with AD-related phenotypes

To examine whether our PRS associates with clinical characteristics, we next investigated the correlation between the PRS.noAPOE and AD-related phenotypes, namely CSF biomarker data and FDG and PiB PET brain imaging data. We performed linear regression model analyses based on three models controlling for seven covariates: age at baseline examination, sex, years of education, the first two PCs, and the doses of *APOE* ε4 and ε2 alleles. Model 1 controlled only age at baseline examination, sex, years of education, and the first two PCs. Models 2 and 3 took into the dose of *APOE* ε4 allele in addition to Model 1. Model 3 also added the dose of *APOE* ε2 allele as a full model.

The CSF tTau/Aβ42 and pTau/Aβ42 ratios were significantly associated with the PRS.noAPOE values. These associations were basically maintained in all models (FDR < 0.05, Wald test; **Table 4a and** **Figure 2**) and reflected the influences of tTau and pTau levels but not Aβ42 levels (**Table S6**).

**Figure 2.**
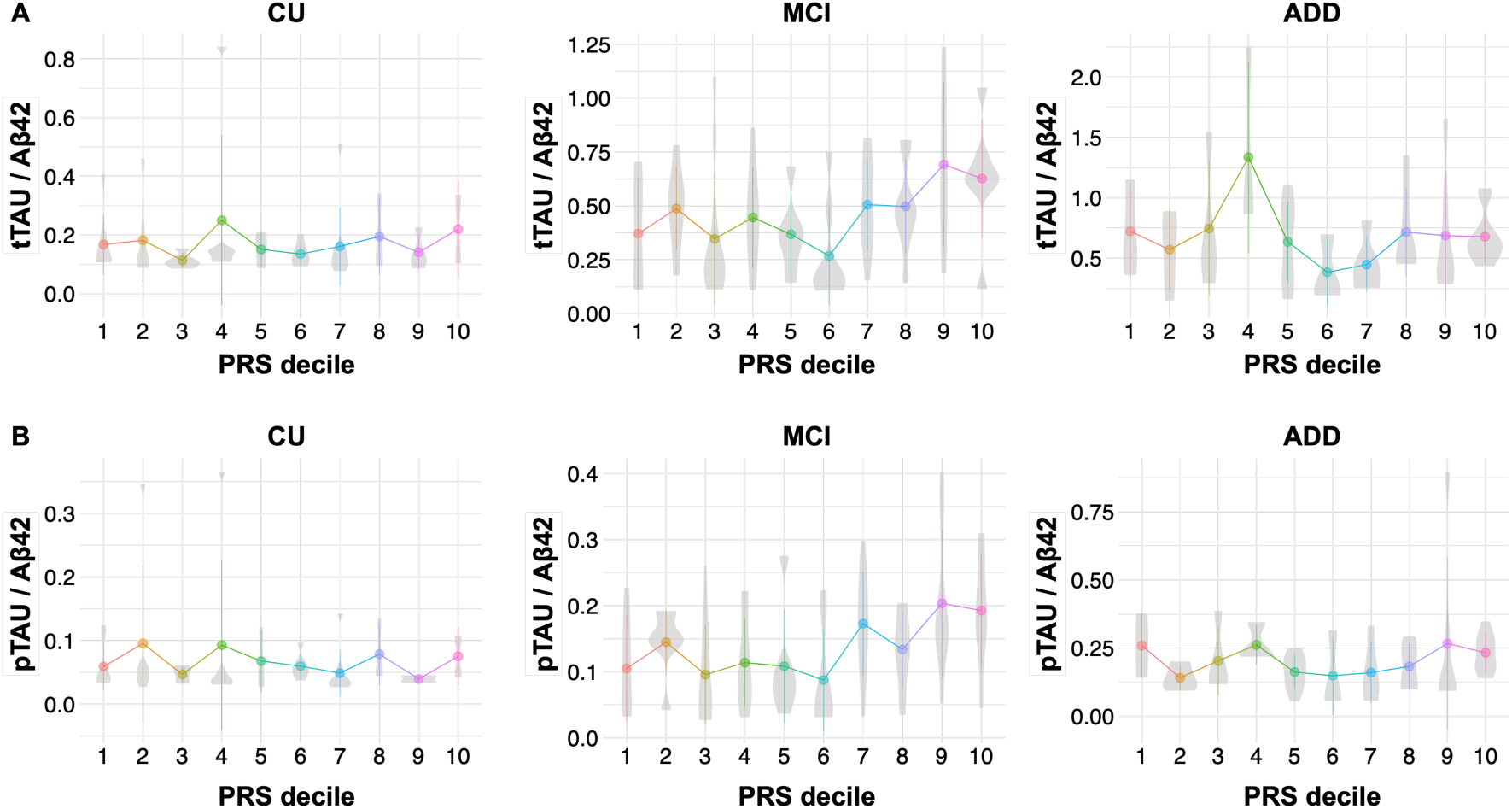
The PRS.noAPOE correlated with CSF Tau/Aβ42 ratios in the MCI. CSF tTau/Aβ42 (A) and pTau/Aβ42 (B) ratios by decile of PRS are shown in each diagnostic group. The participants were divided into ten groups based on the PRS.noAPOE, ranging from the lowest group (1^st^ decile) to the highest group (10^th^ decile). CN = cognitively normal; MCI = mild cognitive impairment; ADD = Alzheimer’s disease dementia.

To investigate the PRS effects to brain atrophy, we first tested the associations between the PRS and the volumes of the entorhinal cortex and hippocampus. Hippocampal volume showed a significant association with the PRS.noAPOE in Model 1 that did not include *APOE* alleles, but this association did not remain significance after FDR correction (p value = 0.042, Wald test; **Table 4b**). We investigated whether the PRS.noAPOE contributes to the discrimination between the normal pattern (N1 pattern) and the AD pattern (P1 pattern) in FDG PET imaging and between negative and positive amyloid scans in PiB PET imaging. As a result, the PRS was associated only with PiB PET imaging (FDR < 0.05, Wald test; **Table 4c**).

We also investigated the correlations between the PRS and cognitive functions. The neuropsychological tests, including the ADAS-Cog, CDR-SB, FAQ, and MMSE, were significantly associated in all models (FDR < 0.01, Wald test; **Table 4d**).

We next stratified the participants into the CU, MCI and ADD groups and examined the association between the PRS.noAPOE and each phenotype. Significant positive correlations between the PRS.noAPOE and CSF tTau/Aβ and between the PRS.noAPOE and pTau/Aβ42 ratios were observed in only the MCI participants (FDR < 0.05, Wald test; **Table 4a**; **Figure 2**). In contrast, these ratios remained stable or reached a plateau relative to the PRS.noAPOE in the CU and ADD participants (**Figure 2**), suggesting that the polygenic burden beyond *APOE* explains some of the heterogeneity in MCI, especially in terms of tau-related biomarker.

### 3.6 *APOE* ε4 non-carriers with high PRS are at high risk of AD conversion

Finally, we examined difference in conversion to AD in the participants with MCI stratified by PRS.noAPOE. We divided MCI participants into three groups based on the PRS.noAPOE distribution of all participants. We compared the conversion to AD of MCI participants in the 1st tertile, referred to as the low-PRS group, and of MCI participants in the 3rd tertile, noted as the high-PRS group. We performed Cox proportional hazard model analysis controlling seven covariates: age at baseline examination, sex, years of education, the first two PCs, and the doses of *APOE* ε4 and ε2 alleles. We did not find significantly different conversion patterns between the high- and low-PRS groups (p value = 0.202, log-rank test; **Table 5 and** **Figure 3**).

**Figure 3.**
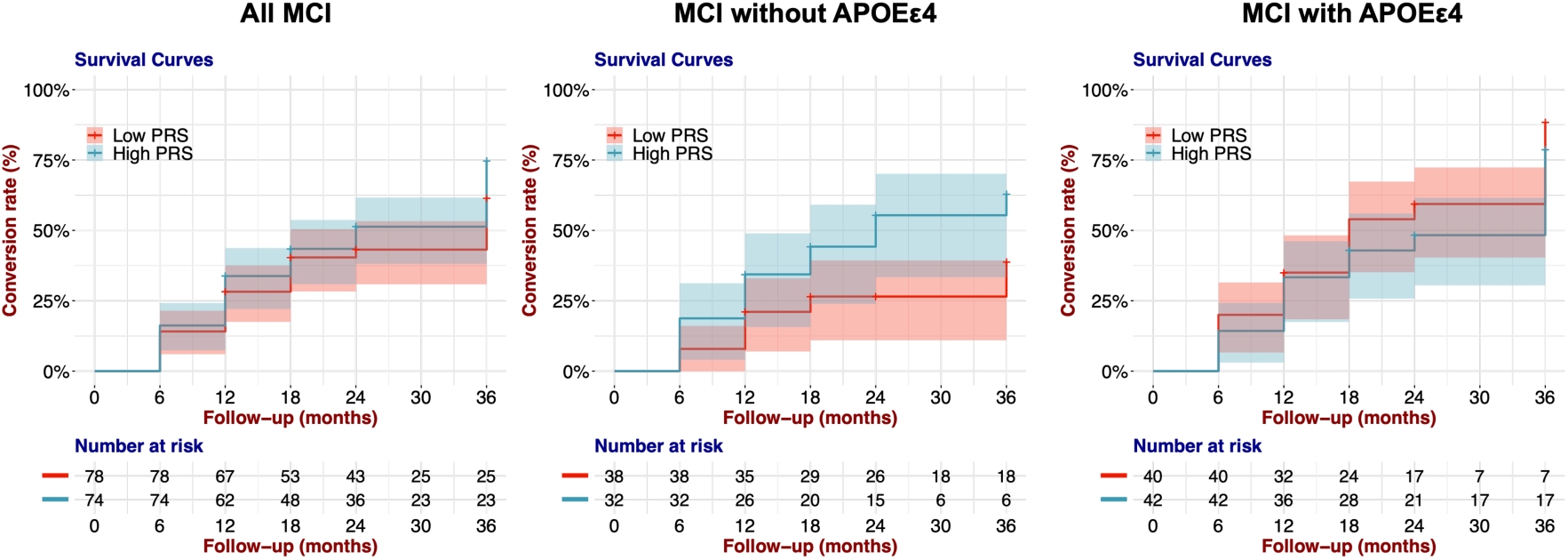
The high-PRS group was more likely to convert to AD than the low-PRS group in the APOE ε4 non-carrier individuals with MCI. Kaplan–Meier survival curves for conversion rates of MCI to AD in the low-PRS group (1^st^ tertile) and the high-PRS group (3^rd^ tertile). The shaded area represents the 95% confidence interval.

When we examined the contribution of each variable, we found that the dose of the *APOE* ε4 allele significantly affected the conversion to AD (HR = 1.604; 95% CI = 1.153-2.230; p value = 0.005, Wald test; **Table 5**), suggesting that this difference in conversion between the two PRS groups was influenced by the *APOE* ε4 allele dose. Therefore, we stratified MCI participants into those with and without *APOE* ε4. In that analysis, we found that among MCI participants without *APOE* ε4, the high-PRS group showed a significantly higher convertion to AD than the low-PRS group (p value = 0.031, log-rank test; **Table 5 and** **Figure 3**). Moreover, the PRS.noAPOE significantly contributed to the difference in AD conversion between the two groups (HR = 2.216; 95% CI = 1.058-4.643; p value = 0.035, Wald test; **Table 5**). We also found no difference in AD conversion among MCI participants with *APOE* ε4 (p value = 0.292, log-rank test; **Table 5 and** **Figure 3**). These results suggested that polygenic effects increase the risk of AD conversion, particularly in MCI subjects without *APOE* ε4.

On the other hand, in *APOE* ε4 carriers, a single factor, namely, *APOE* ε4, may explain much of the AD conversion risk. As expected, there was no significant difference between the *APOE* ε4 noncarrier group with high-PRS and the *APOE* ε4 carrier group (p value = 0.595, log-rank test; **Figure S3**). Although age differences between the groups compared in the above analysis could have affected the results, there were no differences in age at baseline examination between the low- and high-PRS groups or between the converted and nonconverted participants (p value > 0.05, Wilcoxon rank-sum test; **Figure S4**). These results suggest that the PRS contributes to the conversion to AD in participants without *APOE* ε4.

## 4. Discussion

In this study, we evaluated the utility of the PRS for AD in a Japanese cohort. The results showed that the PRS had an effect independent of *APOE* and showed relatively high predictive accuracy when combined with *APOE* ε4. In addition, this effect was replicated in the cohort with a neuropathological diagnosis and the protocol-harmonized independent NA-ADNI cohort. The PRS was significantly associated with CSF tau levels in MCI participants, and MCI with a high PRS was associated with an elevated risk of AD conversion in *APOE* ε4 noncarriers.

Despite the difference in genetic structure between the European and Japanese populations [38], the PRS developed in this study, PRS.noAPOE, showed meaningful predictive accuracy. Such predictive accuracy may be achieved because all participants were diagnosed according to unified inclusion and exclusion criteria and harmonized standardized diagnostic criteria using the same neuropsychological tests (MMSE, CDR-SB, and Wechsler Memory Scale Logical Memory II). The optimal p value threshold for the PRS excluding the *APOE* region was also similar to that reported in previous studies, *p_T_* < 1×10^-5^ [5, 10, 39]. Moreover, while dozens of SNPs were incorporated into these previous PRSs, 131 SNPs were included to calculate the PRS in our study. This difference in the number of SNPs is likely due to differences in genetic structure such as LD blocks. Hence, even if there are racial differences, adding a few dozen SNPs may preserve accuracy.

There is no consensus on the number of SNPs that should be included in the AD PRS. According to a systematic review of PRS studies in AD, PRSs of AD can be organized into two groups: PRSs containing relatively large numbers of SNPs, ranging from 4,431 to 359,500, and PRSs containing relatively small numbers, ranging from 5 to 31 [40]. The latter group is referred to as the oligogenic effect, in contrast to the polygenic effect [41]. From this perspective, our PRS apparently represents an oligogenic effect. Notably, a relatively small number of SNPs has the advantage of providing an inexpensive gene panel. In addition, a PRS composed of many SNPs may be sensitive to geographic differences in genetic structure, whereas a PRS composed of a few dozen SNPs is robust to population bias [42, 43]. However, we should note that our PRS may reflect ancestral differences due to the use of European GWAS statistics. In the future, more robust polygenic effects could be verified by using GWAS statistics for large groups of East Asians, including Japanese individuals.

In our study, 97 genes contributing to the PRS.noAPOE were associated with APP degradation, immunity, and glial cell proliferation. Genetic variants found in a recent AD GWAS were associated with the APP catabolic process and tau protein binding [44]. In addition, many of the genes affected by their genetic variants are expressed in microglia [44]. An analysis of cognitively healthy centenarians in addition to ADD patients and healthy controls revealed that the PRS associated with the immune system was lower in the centenarian group independent of *APOE* ε4, indicating that immune system function is involved in AD resistance [45]. Therefore, our results suggest that common factors related to AD may be shared in the vulnerability of clearance mechanisms and neuroimmune surveillance in the brain among different population.

In our study, the PRS.noAPOE showed significant correlations with CSF tTau/Aβ42 and pTau/Aβ42 ratios only in individuals with MCI. Tau but not Aβ42 strongly influenced this result even controlling *APOE* effect. This correlation may have been observed on in the MCI group because individuals with MCI can have a broad spectrum of clinical characteristics, including CSF tau values, as observed in this study. Interestingly, NA-ADNI studies have shown that the PRS is associated beyond *APOE* with CSF tau but not CSF Aβ42 [43, 46]. From the above, independent studies in different ancestry groups have confirmed that polygenic effects are associated with tau-related biomarkers, especially in individuals with MCI.

Although our results are noteworthy, we must approach the clinical application of our PRS with caution at this stage because the predictive accuracy of our PRS alone is not very high. Similar to currently available PRSs, few biomarkers can perfectly distinguish disease or not; most markers bear some uncertainty. AD and MCI are explained not only by genetic aspects such as PRS, but also by anatomic aspects such as MRI and PET imaging and biological aspects such as CSF biomarkers [47], suggesting that combining multiple biomarkers could compensate for each other’s weaknesses in predictive performance. PRS will allow individuals’ disease risk to be assessed at a relatively early stage, leading to future lifestyle modification and disease prevention.

There were several limitations to this study. First, the CU participants included in the J-ADNI were relatively young. We acknowledge that these CU participants include potential patients who will develop AD in the future. Considering the average age of onset of AD and the allele frequency of *APOE* ε4 in the Japanese population, future work should ideally include CU participants that are over 70 years old [48]. Second, because the number of participants available for the study was small, there was limited power to identify relationships between the PRS and some phenotypes. Larger studies are needed to validate the results of this study. Therefore, combining samples from multiple East Asian cohorts, including cohorts from Japan, is necessary for analysis.

## 5. Conclusion

This study demonstrated that the AD PRS showed a relatively high performance in the Japanese population, despite differences in genetic structure from the European population. Furthermore, this PRS was replicated in the independent Japanese and European cohorts. The AD PRS correlated with phenotypes such as CSF tau levels in MCI. The AD PRS predicted the development of AD in MCI participants without *APOE* ε4. The application of the PRS will allow us to know an individuals’ disease risk at a relatively early life stage, which may lead to future lifestyle modification and disease prevention.

## Abbreviations

(PRS): Polygenic risk score
(AD): Alzheimer’s disease
(J-ADNI): Japanese Alzheimer’s Disease Neuroimaging Initiative
(*APOE*): apolipoprotein E
(Aβ): amyloid-beta
(MCI): mild cognitive impairment
(MRI): magnetic resonance imaging
(CU): cognitively unimpaired
(ADD): Alzheimer’s disease dementia
(NA-ADNI): North American Alzheimer’s Disease Neuroimaging Initiative
(C+T): clumping and thresholding
(AUC): area under the receiver operator characteristic curve
(MDS): multidimensional scaling
(CSF): cerebrospinal fluid
(tTau): total tau
(pTau): phosphorylated tau
(PET): positron emission tomography
(FDG): ^18^F-2-fluoro-2-deoxy-D-glucose
(PiB): ^11^C-Pittsburgh compound B
(MMSE): Mini-Mental State Examination
(FAQ): Functional Assessment Questionnaire
(CDR): Clinical Dementia Rating
(CDR-SB): CDR-Sum of Boxes
(ADAS-Cog): AD Assessment Scale-Cognitive Subscale
(FDR): false discovery rate
(HR): hazard ratio
(CI): confidence interval

## Acknowledgements

We thank all the participants and staff of the J-ADNI and NA-ADNI, and the donors and facility staff for providing autopsy brains. The J-ADNI was supported by the following funding sources: the Translational Research Promotion Project from the New Energy and Industrial Technology Development Organization of Japan; Research on Dementia, Health Labor Sciences Research Grant; the Life Science Database Integration Project of Japan Science and Technology Agency; the Research Association of Biotechnology (Astellas Pharma Inc., Bristol-Myers Squibb, Daiichi-Sankyo, Eisai, Eli Lilly and Company, Merck-Banyu, Mitsubishi Tanabe Pharma, Pfizer Inc., Shionogi & Co., Ltd., Sumitomo Dainippon, and Takeda Pharmaceutical Company), Japan; and a grant from an anonymous foundation. The reference genome data used for this research were originally obtained by participants in the Tailor-made Medical Treatment Program (BioBank Japan: BBJ), led by Prof. Michiaki Kubo; these data are available at the website of the NBDC Human Database/the Japan Science and Technology Agency (JST).

## Authors’ contributions

MK: Study design, analysis and interpretation of data, and manuscript draft. AM and NH: Genotyping analysis, interpretation of data, and manuscript revision. YS, SM, AK, HA: Provision of autopsy brains and manuscript revision. KK, KO, SN, RK, TIwatsubo, and AN: Interpretation of data and manuscript revision. TIkeuchi: Study design, interpretation of data, and manuscript draft. All authors read and approved the final manuscript.

## Availability of data and materials

All the J-ADNI data except for the genome data and the reference genome data were obtained from the NBDC Human Database/the Japan Science and Technology Agency (JST) (https://humandbs.biosciencedbc.jp/en/hum0043-v1), (https://humandbs.biosciencedbc.jp/en/hum0014-latest#JGAS000114rp). GWAS statistics were obtained from the Center for Neurogenomics and Cognitive Research (https://ctg.cncr.nl/software/summary_statistics). The J-ADNI genome data are available on request.

## Declarations

### Ethics approval and consent to participate

This study was approved by the ethics committees of the University of Tokyo, Osaka University and Niigata University.

### Consent for publication

Consent for publication has been granted by J-ADNI administrators.

---

**Figure S1.**
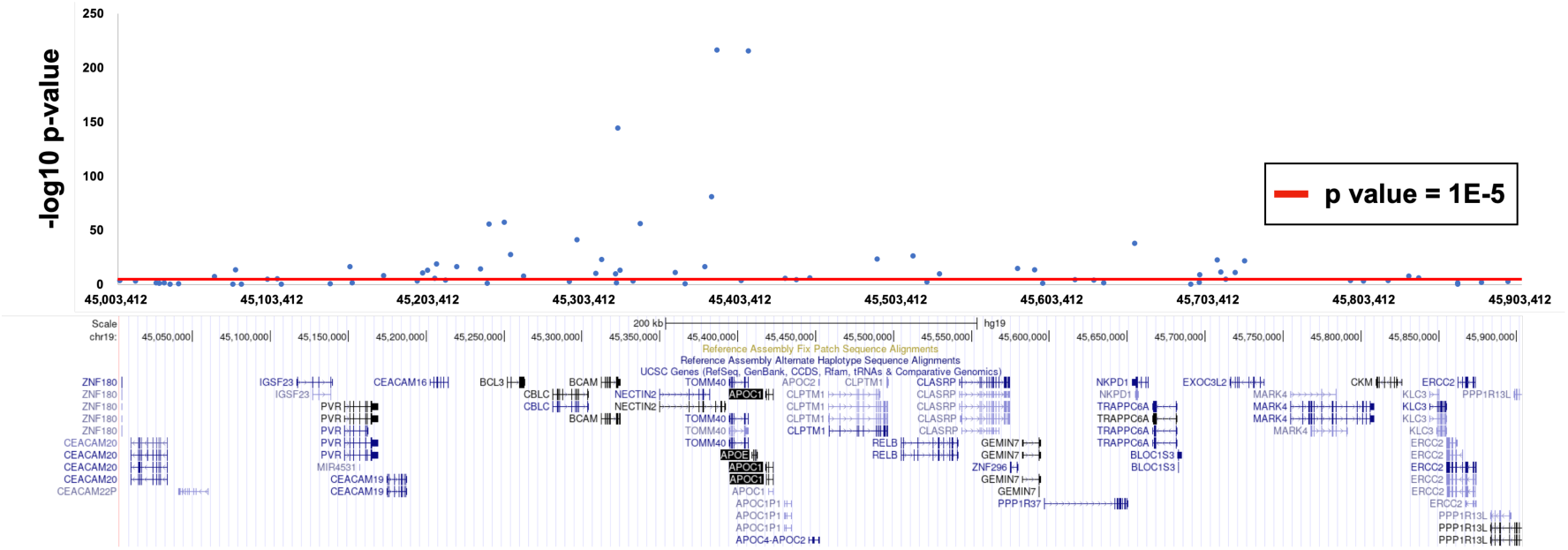
The excluded region around the *APOE* gene. We removed the *APOE* region, consisting of ±500 kb, from around the top-hit SNP rs1160985 (chr19:45403412) in our data. Each data point indicates GWAS p values from Jansen *et al.* [32] used as SNP weights in the PRS calculation.

**Figure S2.**
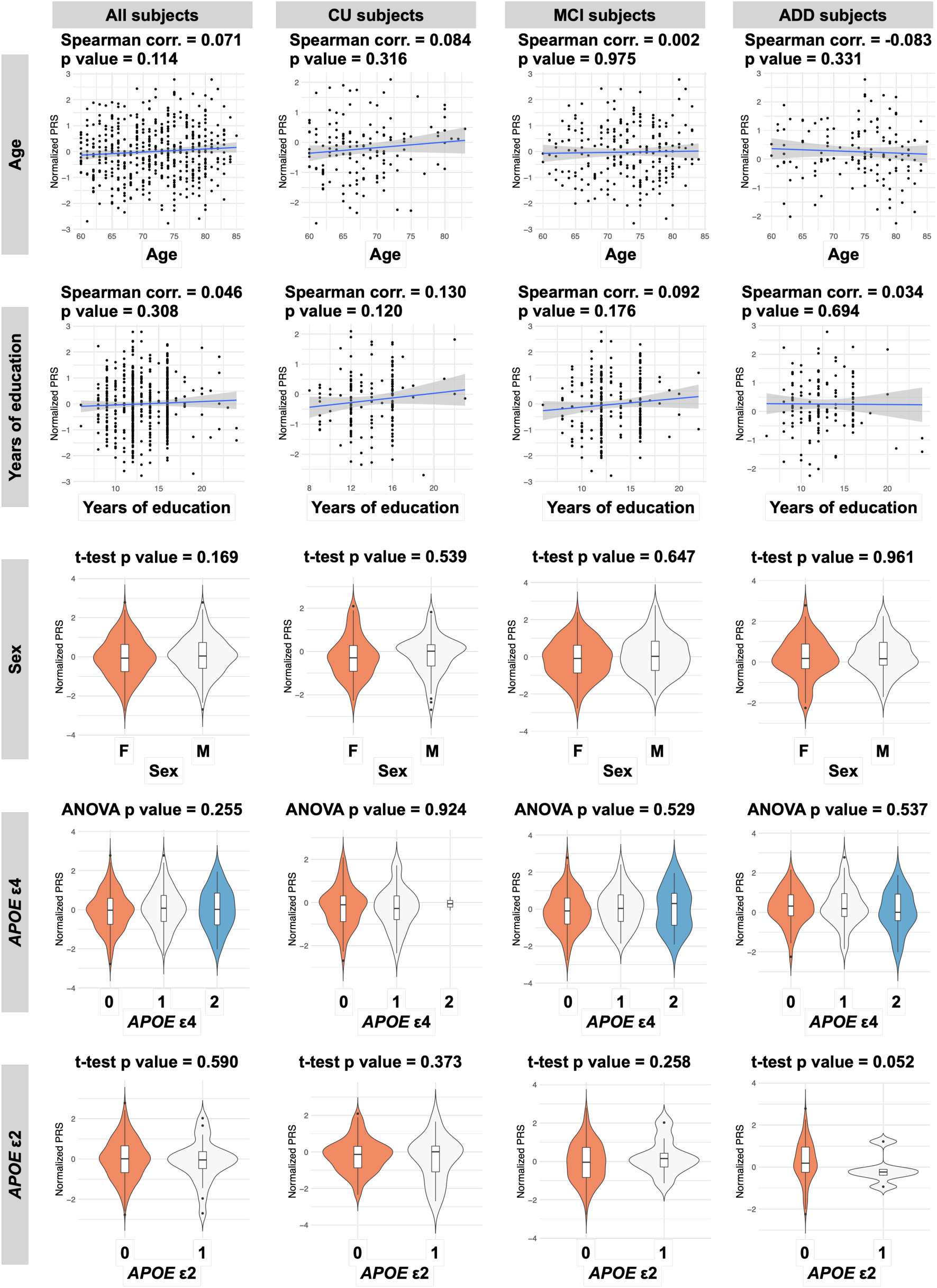
Associations between the PRS and covariates. Age at baseline examination and years of education were examined by Spearman correlation. Sex and doses of *APOE* ε4 and ε2 alleles were analysed by t tests or ANOVAs. CN = cognitively normal; MCI = mild cognitive impairment; ADD = Alzheimer’s disease dementia.

**Figure S3.**
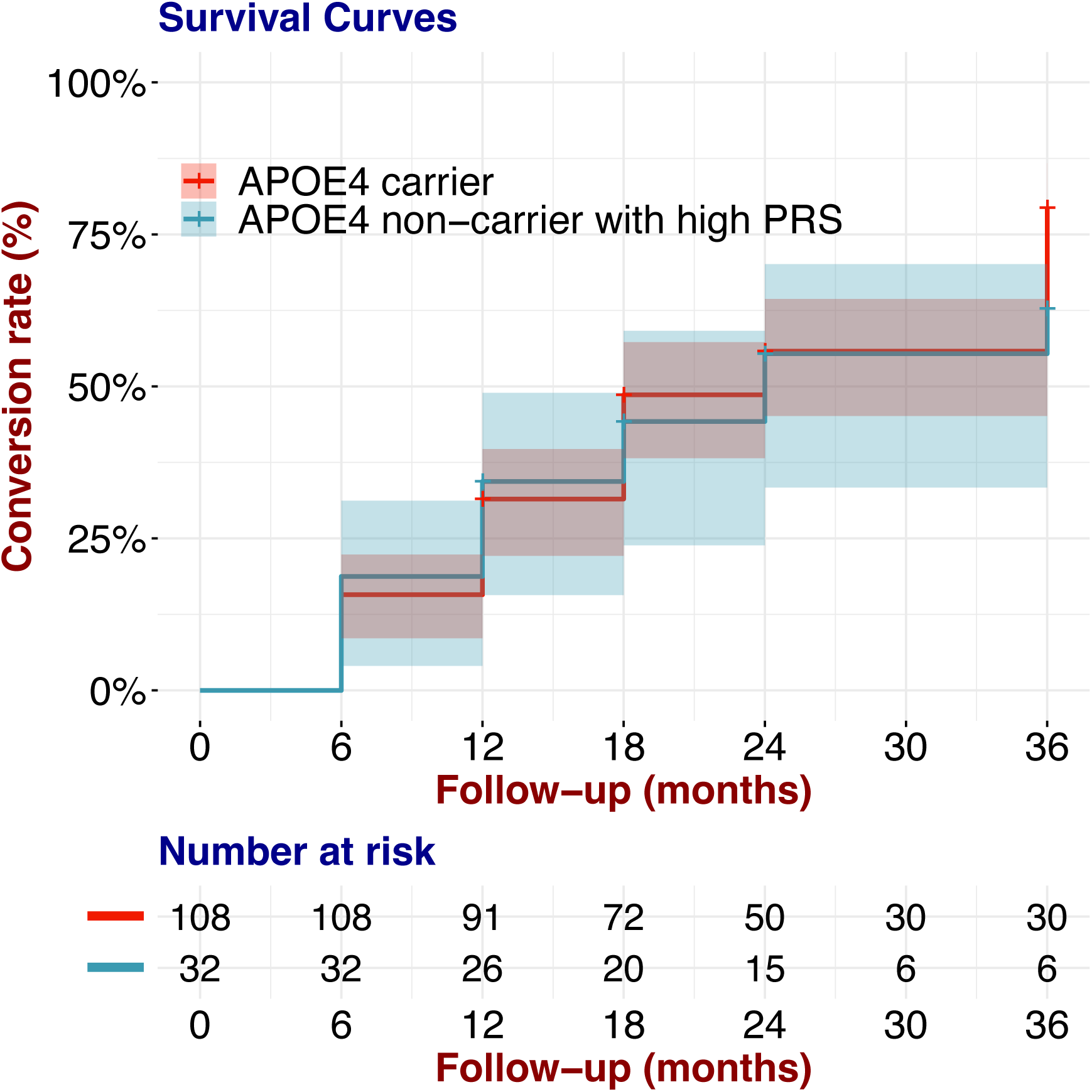
Comparison of AD conversion between *APOE* ε4 carriers and *APOE* ε4 non-carriers with high PRS values. Kaplan–Meier survival curves for the rates of conversion from MCI to AD in *APOE* ε4 carriers and *APOE* ε4 non-carriers with high PRS values. The shaded area represents the 95% confidence interval.

**Figure S4.**
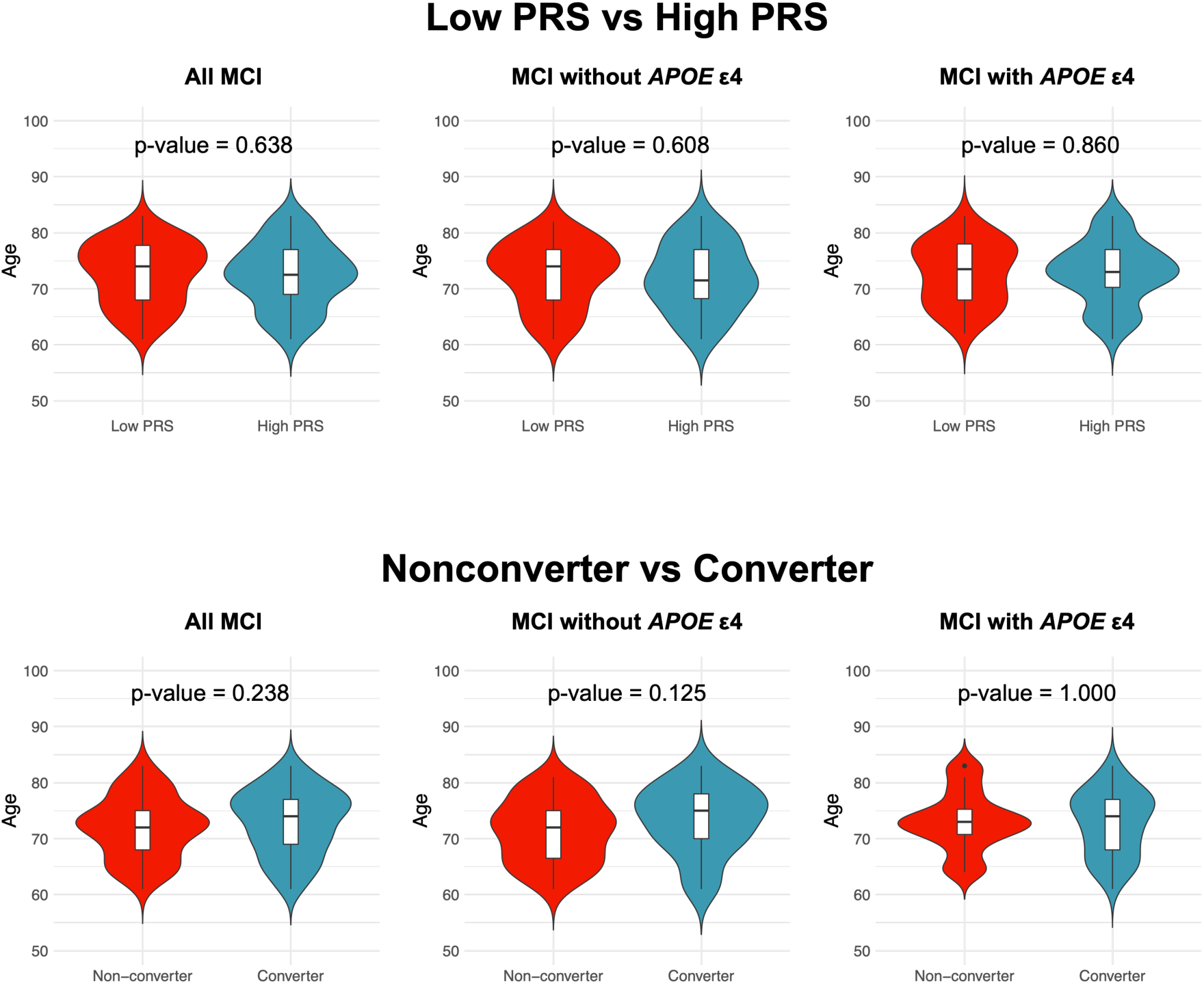
Age differences between the low- and high-PRS groups and between the nonconverters and converters. Baseline ages were compared between groups using the Wilcoxon rank-sum test. Each violin plot includes the kernel probability density of the data at different values and the box plots with the median value and the interquartile range.

